# Maternal per- and poly-fluoroalkyl substances exposures associated with higher depressive symptom scores among immigrant women in the Chemicals in Our Bodies cohort in San Francisco

**DOI:** 10.1101/2022.10.04.22280679

**Authors:** Max T. Aung, Stephanie M. Eick, Amy M. Padula, Sabrina Smith, June-Soo Park, Erin DeMicco, Tracey J. Woodruff, Rachel Morello-Frosch

## Abstract

**Background:** Exposure to per- and poly-fluoroalkyl substances (PFAS) remains an important public health issue due to their widespread detection and persistence in environmental media, slow metabolism in humans, and influences physiological processes such as neurological signaling. Maternal depression is highly prevalent during pregnancy and the postpartum period and is an important neurological outcome that is potentially sensitive to PFAS. The health risks associated with PFAS may be further amplified in historically marginalized communities, including immigrants.

**Objective:** We evaluated the extent to which maternal concentrations of PFAS were associated with depression scores during pregnancy and whether effects differed between US born and immigrant women.

**Methods:** Our analytical sample included 282 US born and 235 immigrant pregnant women enrolled in the Chemicals in Our Bodies prospective birth cohort based in San Francisco, CA. We measured 12 PFAS in serum samples collected in the second trimester and depressive symptom scores were assessed using the Center for Epidemiologic Studies Depression Scale in the same period. Associations were estimated using multiple linear regression, adjusting for maternal age, education, pre-pregnancy body mass index, and parity. Associations with a PFAS mixture were estimated using quantile g-computation.

**Results:** In adjusted linear regression models, a natural log unit increase in two PFAS was associated with higher depression scores in the overall sample, and this association persisted only among immigrant women (*β*[95% confidence interval]: perfluorooctane sulfonic acid (1.3 [0.3-2.3]) and methyl-perfluorooctane sulfonamide acetic acid (1.5 [0.6-2.3]). Using quantile g-computation, we observed that simultaneously increasing all PFAS in the mixture by one quartile was associated with increased depressive symptoms among immigrant women (mean change per quartile increase= 1.12 [0.002, 2.3]), and associations were stronger compared to US born women (mean change per quartile increase= 0.09 [-1.0, 0.8]).

**Conclusions:** Findings provide new evidence that PFAS are associated with higher depression symptoms among immigrant women during pregnancy. Results can inform efforts to address environmental factors that may affect depression among US immigrants.

## 1. Introduction

Endocrine disrupting chemicals (EDCs) such as per- and poly-fluoroalkyl substances (PFAS) are major environmental contaminants of health concern due to their increasing ubiquity and persistence in environmental media, including drinking water (Sunderland et al., 2019). Biomonitoring studies indicate that widespread environmental contamination from consumer products and historic manufacturing has led to detection of multiple PFAS in nearly all humans and wildlife (Calafat et al., 2007; De Silva et al., 2021; Sunderland et al., 2019). While regulations and phaseouts have led to reductions of some PFAS in humans (Hurley et al., 2018), given the long half-life of many of these compounds, there is a need to concurrently determine health risks associated with multiple PFAS exposures to better inform human health risk assessments.

There is increasing evidence that human exposure to PFAS affects several intermediate biological mechanisms, including inflammation, oxidative stress, hormone signaling, and lipid metabolism (Fenton et al., 2021). While PFAS can accumulate in various tissues in the human body, a recent review highlighted evidence from human, animal, and *in vitro* studies indicating that PFAS can accumulate in the brain and nervous tissue potentially by interfering with tight junctions at the blood brain barrier and interacting with transmembrane transporters (Cao and Ng, 2021). Importantly, PFAS may elicit neurotoxic effects through various mechanisms, including disrupting calcium dependent signaling and interfering with neurotransmitter signaling (Cao and Ng, 2021).

Perinatal mood and anxiety disorders occur among 1 in 7 women in the United States (US) during pregnancy and in the postpartum period (Luca et al., 2019, Dagher et al., 2021). Pre- and perinatal depression has downstream implications for adverse outcomes, including increased risk for maternal and perinatal morbidity (*e*.*g*., preterm birth, postnatal depression) (Dagher et al., 2021). The economic costs of mood and anxiety disorders during pregnancy and postpartum, including depression, has been estimated to be $14.2 billion (Luca et al., 2019). Both social and environmental factors can influence the onset and severity of depression (Mutic et al., 2021). For example, structural racism and perceived discrimination may contribute to disparities in depression rates and severity (Hankerson et al., 2022). Furthermore, immigration status may intersect with structural racism and contribute to higher depression rates among immigrant women (Alegría et al., 2017; O’Mahony et al., 2013; Snow et al., 2021). Thus, it is critical to advance understanding of risk factors of depression with close attention towards marginalized immigrant communities.

In addition to understanding social factors that influence perinatal depression disparities, there is a need to determine the contribution of potential environmental neurotoxicants such PFAS. Despite evidence of the toxic effects of PFAS, no studies have assessed the relationship between PFAS and depression, particularly among pregnant women. To address this knowledge gap, we assessed relationships between prenatal PFAS exposures and maternal depressive symptoms during pregnancy. Furthermore, we sought to determine the extent to which these associations differed between US born and immigrant women. We hypothesized that PFAS exposure increases risk of prenatal depression, and that this relationship is amplified among immigrant women partly due to structural factors and social stressors that are unique to immigrants.

## 2. Methods

### Study Population

Participants in this study are part of the Chemicals in Our Bodies (CIOB) prospective pregnancy cohort based in San Francisco, CA. A detailed description of the study population is provided elsewhere (Eick et al., 2021, 2020). Briefly, recruitment of study participants occurred during the second trimester of pregnancy at three University of California, San Francisco (UCSF) hospitals. The inclusion and eligibility criteria for participation included being at least 18 years of age, able to speak English or Spanish as a primary language and having a singleton pregnancy. All study participants provided written, informed consent prior to participation, and the Institutional Review Boards at UCSF (13-12160) and the University of California, Berkeley (2010-05-04) approved the CIOB study.

### PFAS exposure assessment

Serum was collected during the second trimester (range of 12-28 weeks’ gestation) and were stored at - 80° C. Serum samples were analyzed for PFAS at the Environmental Chemical Laboratory at the California Department of Toxic Substances Control (DTSC). Samples were injected into an automated on-line solid phase extraction system coupled to liquid chromatography and tandem mass spectrometry using methods previously described in detail (Eick et al., 2021, 2020). Twelve unique PFAS compounds were measured: perfluorobutane sulfonate (PFBS), perfluorohexanesulphonic acid (PFHxS), perfluorooctane sulfonic acid (PFOS), perfluoroheptanoic acid (PFHpA), perfluorooctanoic acid (PFOA), perfluorononanoic acid (PFNA), perfluorodecanoic acid (PFDeA), perfluoroundecanoic acid (PFUdA), perfluorododecanoic acid (PFDoA), perfluorooxtane sulfonamide (PFOSA), methyl-perfluorooxtane sulfonamide acetic acid (Me-PFOSA-AcOH), and ethyl-perfluorooctane sulfonamide acetic acid (Et-PFOSA-AcOH). Among these quantified compounds, we focused downstream analyses on those with at least 70% detection rates, which led to a final PFAS roster of PFOA, PFOS, PFNA, PFHxS, PFDeA, PFUdA, and Me-PFOSA-AcOH. For any values that were below the method detection limit (MDL), we imputed the machine read value if it was available. We chose this imputation method in order to maximize all available data and preserve a more realistic exposure distribution, as setting all minimally detected values to the same value in reality is implausible. When machine read values were not available, we imputed using the MDL/√2.

### Depression symptom score assessment

During the second trimester, we administered the Center for Epidemiological Studies-Depression (CES-D), a validated, self-report instrument, to assess depressive symptoms. The CES-D collects participants’ responses to a 20-item questionnaire to measure symptoms of depression that have occurred in the prior year, which includes factors such as feeling unusually bothered, having poor appetite, feeling hopelessness, trouble focusing, feeling fearful, trouble sleeping, feeling lonely, pessimism, and fatigue (Radloff, 1977). While the total CES-D score can be set to a binary value at or above 16 to designate depression, we selected to use this binary categorization only for descriptive statistics and modeled the continuous CES-D score for all regression models. The rationale for this was that modeling continuous CES-D score allows for estimation of the relative increase in depressive symptoms and is sufficiently powered for models with limited sample size.

### Statistical approach

We estimated univariate distributions of PFAS chemicals in our overall sample and stratified by immigrant status and estimated Spearman correlation coefficients between compounds. We also estimated PFAS concentrations and tabulated demographic and social factors between participants with likely depression and no depression (<u>>16</u> vs. <16). We tested for bivariate associations using t-tests and chi-squared tests, for continuous and categorical variables, respectively.

For single pollutant analyses, we built multiple linear regression models with CES-D scores as a continuous outcome and individual natural log transformed PFAS metabolites as independent exposures. For adjusted models, we considered potential confounders and determined final covariates based on bivariate associations, which included: maternal age, education, pre-pregnancy body mass index (BMI), parity, and immigrant status. While single time point BMI or change in BMI during pregnancy may be a mediator for PFAS and CES-D score measures during pregnancy, we conceptualized pre-pregnancy BMI to serve as a proxy for dietary and physical behavior characteristics prior to pregnancy, which would be temporarily and biologically consistent with a confounder for PFAS and CES-D scores measured during pregnancy. We then stratified our sample based on immigrant status, therefore including all covariates except immigrant status. We observed evidence of non-linearity in CES-D scores, and we explored transformations [ln(CES-D+1)] as potential sensitivity analyses, and based on comparable residual distributions and greater interpretability, we modeled CES-D scores without transformation in final models.

For mixtures analyses, we applied quantile g-computation, which estimates the cumulative effect of simultaneously increasing all PFAS by one quartile within the mixture using a parametric, generalized linear based implementation of g-computation (Keil et al., 2020). With this approach, individual PFAS are allowed to have opposing directions of effect relative to CES-D scores, which are translated to negative and positive weights that sum to 1 and are indicative of relative compound importance either in the negative or positive directions.

## 3. Results

**Table 1** reports key covariates relative to CES-D score cut-off of 16 for individuals with symptoms indicating risk for clinical depression (N=42 participants [8.1%]). The CIOB study sample was comprised largely of White (42.2%) and Latina (32.4%) participants. Approximately 56% of participants were US born and 44% were immigrants. Half of the study participants reported that the current pregnancy was their first. Most participants (65.2%) had college-level educational attainment. Most participants reported having no history of smoking (95.3%). Each of these covariates were associated with risk of depression (**Table 1**). For example, risk of depression was highest among Latina participants (66.7%), women who did not graduate college (82.5%), and immigrant participants (63.2%).

**Table 1.**
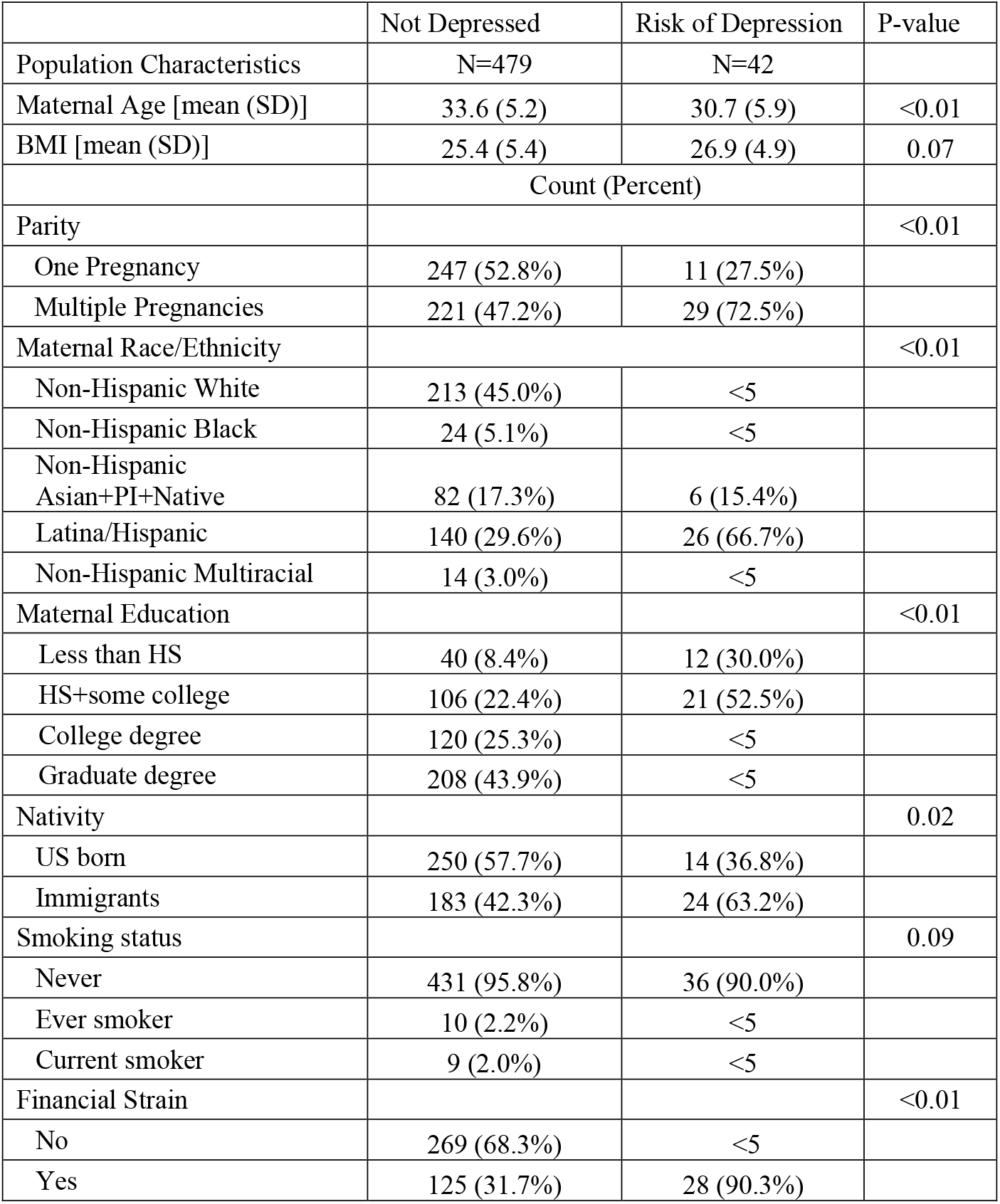
Chemicals in Our Bodies cohort descriptive statistics by depression indicator.

The crude distributions of two PFAS compounds differed by depression status (**Table 2**). PFHxS and PFUdA were both lower in those with risk of depression (Geometric mean [GM_PFHxS_] = 0.27 ng/mL; GM_PFUdA_ = 0.05 ng/mL) compared to those without risk of depression (GM_PFHxS_ = 0.37 ng/mL; GM_PFUdA_= 0.1 ng/mL). PFAS compounds were all positively correlated, and we observed the smallest positive correlation between PFUdA and MePFOS (ρ=0.1) and the largest positive correlation between PFDeA and PFNA (ρ= 0.7) (**Supplemental Table 1**). Stratification by immigrant status also indicated that all PFAS were lower among immigrants compared to US born participants. The greatest difference was observed for PFOS, where US born women had a GM of 2.2 ng/mL, while immigrant women had a GM of 1.5 ng/mL.

**Table 2.** PFAS distributions in relation to depression risk and nativity status

|  |  | Not Depressed | Risk of depression |  | US born | Immigrants |  |
| --- | --- | --- | --- | --- | --- | --- | --- |
| PFAS compounds<br>(ng/mL) [median (25<sup>th</sup><br>and 75<sup>th</sup> percentile)] | MDL<sup>1</sup> | N=479 | N=42 | P-value | N=282 | N=235 | P-value |
| PFNA | [0.04, 0.06] | 0.3 [0.2; 0.4] | 0.3 [0.2; 0.4] | 0.19 | 0.3 [0.2; 0.5] | 0.2 [0.2; 0.4] | <0.01 |
| PFOA | [0.06, 0.13] | 0.8 [0.5; 1.2] | 0.7 [0.5; 0.9] | 0.21 | 0.9 [0.6; 1.3] | 0.6 [0.4; 0.9] | <0.01 |
| PFHxS | [0.01, 0.02] | 0.3 [0.2; 0.6] | 0.3 [0.2; 0.4] | 0.02 | 0.4 [0.3; 0.7] | 0.2 [0.2; 0.4] | <0.01 |
| PFOS | [0.06, 0.07] | 2.0 [1.3; 3.1] | 1.6 [1.05; 2.6] | 0.15 | 2.2 [1.5; 3.3] | 1.5 [1.0; 2.7] | <0.01 |
| Me-PFOSA-AcOH | [0.01] | 0.04 [0.03; 0.07] | 0.05 [0.04; 0.08] | 0.04 | 0.05 [0.03; 0.08] | 0.04 [0.02; 0.05] | <0.01 |
| PFDeA | [0.06, 0.08] | 0.1 [0.08; 0.2] | 0.08 [0.06; 0.2] | 0.03 | 0.1 [0.08; 0.2] | 0.1 [0.07; 0.2] | 0.09 |
| PFUdA | [0.03, 0.05] | 0.1 [0.06; 0.2] | 0.06 [0.04; 0.09] | <0.01 | 0.1 [0.07; 0.2] | 0.08 [0.04; 0.2] | <0.01 |

Abbreviations: per- and poly-fluoroalkyl substances (PFAS), perfluorononanoic acid (PFNA), perfluorooctanoic acid (PFOA), perfluorohexanesulphonic acid (PFHxS), perfluorooctane sulfonic acid (PFOS), methyl-perfluorooxtane sulfonamide acetic acid (Me-PFOSA-AcOH), perfluorodecanoic acid (PFDeA), perfluoroundecanoic acid (PFUdA), geometric mean (GM), geometric standard deviation (GSD), method detection limit (MDL)
<sup>1</sup> Multiple MDLs are presented where batches had different LODs with the same lab.

We observed associations between single pollutants and PFAS mixtures in relation to depression scores (**Table 3)**. In the overall sample, we observed that a natural log increase in PFOS was associated with 0.8 unit increase in CES-D scores (95% confidence interval [CI]= 0.1, 1.4) and Me-PFOSA-AcOH was associated with a 0.7 unit increase in CES-D scores (95% CI= 0.2, 1.2). When stratifying by immigrant status, we observed that these associations were largely driven and more amplified among immigrant compared to US born women. A natural log unit increase in PFOS in immigrant women was associated with a 1.3 unit increase in CES-D scores (95% CI= 0.3, 2.3), while Me-PFOSA-AcOH was associated with a 1.5 unit increase in CES-D score (95% CI= 0.6, 2.3). Quantile g-computation indicated similar trends, where simultaneously increasing all PFAS in the mixture by one quartile was associated with a greater increase in CES-D scores in immigrant women (mean change per quartile increase= 1.1, 95% CI= 0.002, 2.3) compared to US born women (mean change per quartile increase= -0.09, 95% CI= -1.0, 0.8). Decomposition of quantile g-computation effect estimates indicated that Me-PFOSA-AcOH exhibited the greatest positive weights to the overall effect relative to any of the other PFAS compounds in the overall sample and among immigrant women and US born women (**Supplemental Table 2**).

**Table 3.**
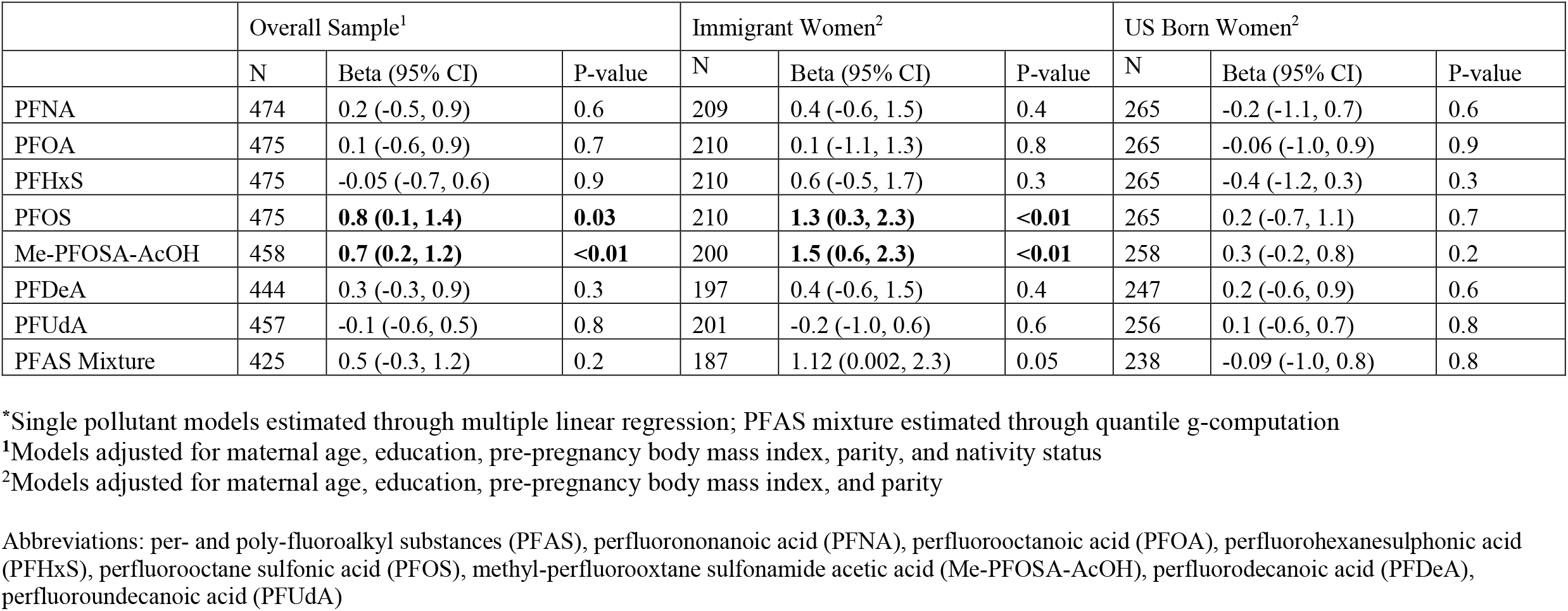
Estimates of association and 95% confidence intervals (CI) between PFAS^*^ (single natural log-transformed compounds and total mixture) and maternal CES-D scores.

## 4. Discussion

In this study we investigated the relationship between maternal prenatal CES-D scores in the CIOB cohort and prenatal serum PFAS concentrations. Importantly we investigated stratified associations based on immigrant status to better understand potential effect modification by this important social identity. Our analysis provides evidence that two long-chained PFAS compounds, PFOS and Me-PFOSA-AcOH, are associated with higher depressive symptom scores, and that this association was more pronounced among immigrant participants. Our mixtures analysis suggests that simultaneous exposure to multiple PFAS was associated with increased CES-D scores. As with single pollutant models, the associations between the overall PFAS mixture and higher depression scores were evident largely among immigrant women and not US born women.

One other study conducted in 2013 has investigated PFAS exposures among immigrants, and this was focused on potential exposures through consumption of fish among refugees and immigrants from Myanmar who resided in Buffalo NY (Liu et al., 2022). Interestingly, the study found immigrants from Myanmar had substantially higher levels of several PFAS, including PFOS (median = 35.6 ng/mL) compared to a sample of licensed anglers (median = 11.6 ng/mL) and the general NHANES population from 2013-2014 (5.6 ng/mL). In contrast, our more recently conducted study found PFAS was lower in all immigrant women compared to US born women, which may partly be driven by historical trends in decreasing PFAS levels due to phase-outs and remediation efforts. The stronger associations we observed between PFOS and Me-PFOSA-AcOH and higher depressive symptom scores among immigrant women suggest that this may be a vulnerable population of concern. Immigrants can experience marginalization, which in turn can influence their exposures to social stressors such as discrimination, social isolation, acculturative stress, financial hardship, and barriers to educational attainment (Gee et al., 2016; Miranda et al., 2005; Zelkowitz et al., 2008). Co-exposure to social stressors alongside PFAS exposures may enhance or amplify depressive symptoms even when there is lower average levels of PFAS in immigrants compared to those that are US born (Morello-Frosch and Shenassa, 2006). Thus, greater attention needs to be given to exposure-response functions for health outcomes in marginalized groups, even when their exposure levels may be lower than national averages.

To our knowledge, no previous studies have investigated the relationships between PFAS and depressive symptoms during pregnancy, although there are important parallels that can be drawn from studies that have focused on neurobehavioral development in children, since this may be an antecedent precursor to psychiatric conditions later in life. For example, the Health Outcomes and Measures of the Environment (HOME) study investigated prenatal and childhood PFAS exposures at ages 3 (n=146) and 8 (n=193) in association with parental-report responses using the Behavioral Assessment System for Children-2 (BASC-2) (Vuong et al., 2021). Although childhood PFAS exposures were not found to be associated with neurobehavioral features in this study, prenatal PFOS exposure was associated with higher BASC-2 scores for externalizing problems, hyperactivity, aggression and conduct problems (Vuong et al., 2021). A separate study based in the Faroe Islands, found prenatal maternal (n=449) PFOS to be modestly associated with lower childhood cognitive scores using the Boston Naming Test (Oulhote et al., 2019). A previous review has also highlighted that overall there are mixed findings, and noted some where null associations were observed between PFAS compounds and some childhood neurobehavioral outcomes such as attention deficit hyperactivity disorder (Liew et al., 2018). A meta-analysis identified evidence that childhood externalizing problems were associated with subsequent adult depression risk (Loth et al., 2014). While direct comparisons from these studies cannot be made with findings from our present study, developmental effects observed in these studies provide biological plausibility that PFAS can affect neurological health and thus potentially adult depression risk.

Neuroinflammation and subsequent impacts on neuronal signaling have been hypothesized as potential precursors in the etiology of depression (Troubat et al., 2021). The potential mechanisms of action linking PFOS and Me-PFOSA-AcOH to neurological outcomes may be driven by neuroinflammation, intracellular calcium levels in affected neurons, and alterations with neurotransmitters (Cao and Ng, 2021). For example, a study of pregnant women (n=725) based in Shanghai, China, determined that prenatal PFAS mixtures was associated elevated brain derived neurotrophic factor (BDNF) in cord blood of male offspring (Yu et al., 2021). BDNF is a peptide largely expressed in the central nervous system that is involved with neuronal development and plasticity, hence elevated cord blood levels may be indicative of neurotoxic effects induced by PFAS (Hao et al., 2017; Yu et al., 2021). Relatedly, animal models have shown evidence that PFOS and PFAS mixtures can affect neurotransmitter concentrations, such as acetylcholine and cholinergic system signaling (Cao and Ng, 2021; Foguth et al., 2020; Johansson et al., 2008). Evidence from animal models also indicates that long-chained PFAS (*e*.*g*., PFOS and Me-PFOSA-AcOH) may cross the blood brain barrier with slightly more efficiency than shorter chained PFAS, and a recent human study indicates that PFAS detection in cerebral spinal fluid correlates with elevated c-reactive protein, indicating a coupled inflammatory relationship with PFAS crossing the blood brain barrier (Cao and Ng, 2021; Dassuncao et al., 2019; Wang et al., 2018). Altogether, these potential mechanisms should be explored in future studies of PFAS and depression to contextualize intermediate physiological states attributable to the associations we observed in our study.

Given that PFAS are endocrine disrupting chemicals, their influence on neurological health may also be driven through hormone disruption. For example, a previous cross-sectional study in NHANES (n=1,886) showed that several PFAS, including PFOS, were associated with higher total and free testosterone, in addition to non-linear associations with sex hormone binding globulin (Xie et al., 2021). Another prospective birth cohort study in Shanghai (n=1,842) reported positive associations between PFAS mixtures and the thyroid hormone free thyroxine as well as a non-linear relationship between PFOS and thyroid stimulating hormone (Aimuzi et al., 2020). Hormone disruption may be an important factor linking PFAS exposures to depression outcomes due to the influence of hormones on neuroinflammation (Slavich and Sacher, 2019), and this relationship should be explored in future studies to contextualize our findings.

Our study had several limitations. First, our indicator of depression was based on self-report and we do not have a clinical confirmed depression outcome measure; however, the CES-D is validated and has been widely used during pregnancy (Heller et al., 2022). Second, our study design was cross-sectional as serum samples and the CES-D were collected at the same second trimester visit, which raises potential temporality issues related to PFAS exposures and depressive symptom outcomes. However, given the long half-life of PFAS, it is unlikely that our results would be subject to reverse causality. In addition, we did not measure other co-morbidities of relevance for depression during pregnancy such as anxiety. Future studies should more comprehensively assess depression and potential co-morbidities such as anxiety during pregnancy as well as during the post-partum period. Another limitation in our study is the absence and insufficient power for data disaggregation among different groups of immigrants. This is an important consideration because evidence from a recent study leveraging the National Health Interview Study indicated that among immigrants, the highest rates of depression were observed among immigrants from Mexico, Central America, or the Caribbean (Flores Morales and Nkimbeng, 2021). We are unable to determine if there are specific immigrant ethnic identities that may experience greater susceptibility to PFAS related depression symptom risk. Nonetheless, our cohort had substantially balanced and high numbers of immigrant women to assess risk of depressive symptoms. Future studies should expand data disaggregation efforts. Despite these limitations, our study contained notable strengths. The CiOB is a well-characterized cohort and is highly diverse both in terms of self-identified race/ethnicity, immigration status, and socioeconomic status. The CiOB cohort has relatively high proportions of Latina and Asian American immigrants, two of the fastest growing immigrant populations in the US (Abby Budiman et al., 2020). Finally, our application of quantile g-computation to assess PFAS mixtures advances greater understanding of cumulative PFAS in association with depressive symptoms risk.

In conclusion, results from our study find that prenatal PFAS exposure is a significant risk factor for depression among immigrants. These findings have important implications for contextualizing unique environmental risk factors among immigrants and can help inform regulatory and health protective policies surrounding immigrants and mental health. Future studies should confirm these findings among other immigrant groups, more comprehensively assess depressive and anxiety symptoms during the prenatal and postpartum periods and elucidate intermediate effect biomarkers and mechanistic pathways for understanding the link between PFAS and depressive symptoms in this uniquely vulnerable population.

## Data Availability

Per University of California, San Francisco Institutional Review Board approval, the data that support the findings of this study are restricted for transmission to those outside the primary investigative team. Data sharing with investigators outside the team requires IRB approval. Requests may be submitted to the Program on Reproductive Health and the Environment (PRHE)

## Acknowledgements

We would like to thank our clinical research coordinators for collecting the data and the data analysis teams for helping to enter and compile the data, particularly Aileen Andrade, Cheryl Godwinde Medina, Cynthia Melgoza Canchola, Tali Felson, Harim Lee, Maribel Juarez, Lynn Harvey and Allison Landowski from the CIOB group. We also thank the study participants who participated in the CIOB cohort. Lastly, we would like to thank the DTSC biomonitoring team (Dr. Hyoung-Gee Baek and Chris Ranque) for the laboratory analysis of PFAS in serum.

## Author Contributions

MTA (Conceptualization, formal analysis, investigation, methodology, writing); SME (Conceptualization, methodology, data curation, reviewing and editing); AMP (Data curation, funding acquisition, reviewing and editing); SS (Formal analysis, data curation, reviewing and editing); JSP (Formal analysis, data curation, reviewing and editing); ED (Data curation, funding acquisition, reviewing and editing); TJW (Funding acquisition, data curation, reviewing and editing); RMF (Conceptualization, methodology, funding acquisition, data curation, reviewing and editing).

