## Supplemental Tables for "Maternal per- and poly-fluoroalkyl substances exposures associated with higher depressive symptom scores among immigrant women in the Chemicals in Our Bodies cohort in San Francisco"

**Supplemental Table 1.** Spearman Correlation Coefficients of PFAS compounds (N=521)

|  | PFNA | PFOA | PFHxS | PFOS | MePFOS | PFDeA | PFUdA |
| --- | --- | --- | --- | --- | --- | --- | --- |
| PFNA | 1 |  |  |  |  |  |  |
| PFOA | 0.7 | 1 |  |  |  |  |  |
| PFHxS | 0.5 | 0.7 | 1 |  |  |  |  |
| PFOS | 0.7 | 0.6 | 0.6 | 1 |  |  |  |
| MePFOS | 0.2 | 0.2 | 0.2 | 0.3 | 1 |  |  |
| PFDeA | 0.7 | 0.6 | 0.3 | 0.6 | 0.1 | 1 |  |
| PFUdA | 0.6 | 0.4 | 0.3 | 0.5 | 0.1 | 0.8 | 1 |

Abbreviations: per- and poly-fluoroalkyl substances (PFAS), perfluorononanoic acid (PFNA), perfluorooctanoic acid (PFOA), perfluorohexanesulphonic acid (PFHxS), perfluorooctane sulfonic acid (PFOS), methyl-perfluorooxtane sulfonamide acetic acid (Me-PFOSA-AcOH), perfluorodecanoic acid (PFDeA), perfluoroundecanoic acid (PFUdA)

**Supplemental Table 2.** Relative positive (+) and negative (-) weights^1^ estimated from quantile q-computation for each PFAS

compound within the overall mixture

| Combined Sample (N=425) | | Immigrant (N=187) | | US Born (N=238) | |
| --- | --- | --- | --- | --- | --- |
| Me-PFOSA-AcOH (+) | 0.43 | Me-PFOSA-AcOH (+) | 0.27 | Me-PFOSA-AcOH (+) | 0.65 |
| PFOS (+) | 0.43 | PFDeA (+) | 0.26 | PFOS (+) | 0.26 |
| PFDeA (+) | 0.14 | PFOS (+) | 0.19 | PFDeA (+) | 0.09 |
| PFUdA (-) | 0.5 | PFHxS (+) | 0.18 | PFNA (-) | 0.48 |
| PFOA (-) | 0.34 | PFNA (+) | 0.1 | PFUdA (-) | 0.23 |
| PFHxS (-) | 0.14 | PFUdA (-) | 0.63 | PFHxS (-) | 0.19 |
| PFNA (-) | 0.008 | PFNA (-) | 0.37 | PFOA (-) | 0.09 |

^1^Positive and negative weights sum to 1

Abbreviations: per- and poly-fluoroalkyl substances (PFAS), perfluorononanoic acid (PFNA), perfluorooctanoic acid (PFOA), perfluorohexanesulphonic acid (PFHxS), perfluorooctane sulfonic acid (PFOS), methyl-perfluorooxtane sulfonamide acetic acid (Me-PFOSA-AcOH), perfluorodecanoic acid (PFDeA), perfluoroundecanoic acid (PFUdA)
